# Method development and characterization of the low molecular weight peptidome of human wound fluids

**DOI:** 10.1101/2020.10.29.20222208

**Authors:** Mariena J.A. van der Plas, Jun Cai, Jitka Petrlova, Karim Saleh, Sven Kjellström, Artur Schmidtchen

## Abstract

Wound infections are significant challenges globally, and there is an unmet need for better diagnosis of wound healing status and infection. The wound healing process is characterized by proteolytic events that are the result of basic physiological processes, but also dysfunctional activations by endogenous and bacterial proteases. Peptides, downstream reporters of these proteolytic actions, could therefore serve as a promising tool for diagnosis of wounds.

Here, we demonstrate a method for the characterisation of the peptidome of wound fluids. We compare acute non-infected wound fluids obtained post-surgery with plasma samples and find significantly higher protein and peptide numbers in wound fluids, which typically were also smaller in size as compared to plasma-derived peptides. Furthermore, we analyse wound fluids collected from dressings after facial skin graft surgery and compare three uninfected, healing wounds with three inflamed *Staphylococcus aureus* infected wounds. The results identify unique peptide patterns of various proteins, including coagulation and complement factors, proteases and antiproteinases.

Together, the work defines a workflow for analysis of peptides derived from wound fluids and demonstrate a proof-of-concept that such fluids can be used for analysis of qualitative differences of peptide patterns from larger patient cohorts, providing potential biomarkers for wound healing and infection.

## Introduction

Wound infections after surgery and in relation to burns, as well as in non-healing wounds are significant medical and societal problems (1). Surgical site infections (SSIs) are leading nosocomial infections in developing countries and the second most frequent nosocomial infections in Europe and the United States (2). For example, in European hospitals, the overall rates of surgical site infection (SSI) range between 3% and 4% of patients undergoing surgery (3). In some procedures, such as when using skin grafts, or performing hernia surgery using biomaterials, the infection risk is higher and may exceed 5-10% (3-5). The economic burden of failing skin repair is therefore extensive (6). Currently, the costs of treating wounds are estimated to be over 3% of the total health care budgets of Western countries (7, 8) and these expenses are projected to grow with the increasing development of antimicrobial resistance, as well as ageing of the population and the rising incidence and prevalence of diseases such as obesity and diabetes (9). All these factors contribute not only to an increased risk for SSI, but also to an increase in the incidence of non-healing wounds in elderly patients with circulatory insufficiencies. Besides the enormous economic burden, wound infections in acute and non-healing wounds lead to increased risk of invasive infections and sepsis, dysfunctional wound closure and risk for unaesthetic scarring, as well as a reduced quality of life (10).

Given the above, there is an unmet need for improved methods to measure and predict wound healing status and infection risk in different types of wounds. Furthermore, detailed characterisation of wounds may enhance our understanding of the causes that lead to failure and possibly reveal novel therapeutic targets. A non-invasive way of investigating wound healing is through analysis of wound exudates. These contain proteins, peptides and other biological components, such as metabolites, which can be characterised to various degrees. In agreement, several studies have reported the use of proteomics for the analysis of wound fluids (11-13). However, although a powerful methodology, mass spectrometry analyses on samples after trypsin digestion of the proteins mainly report on presence of proteins and their high molecular weight fragments, whereas information about the endogenous fragmentation and resulting peptides is lost. In wounds, however, endogenous protein degradation is of high relevance for the understanding of healing as balanced proteolytic activity is essential for progression of healing, whereas aberrant protease activity, such as seen in patients with infected acute wounds (14, 15) and non-healing ulcers (14, 16), may lead to deteriorated wound healing. Various endogenous proteases, such as neutrophil elastase, matrix metalloproteases, and collagenases, play important roles in both functional and impaired healing (14, 17-19). Additionally, exogenous proteases secreted by colonizing or infecting bacteria may influence healing as well (20, 21). These different proteases may degrade endogenous proteins into peptides with sequences that are enzyme specific, and this may result in peptide patterns that reflect the nature and level of protein degradation occurring in a wound. This subject has been addressed to some extend by Sabino et al. who applied quantitative proteomics strategies to assess the wound proteome and the activity of distinct protease groups along the healing process, thus mapping proteolytic pathways (22, 23). However, given the dynamics of wound healing and the highly proteolytic environment, the generation of endogenous peptides in wounds, their structures, and possible utilization as biomarkers for wound healing still remains to be explored.

The large scale analysis of endogenous peptides has since its introduction in 2001 (24-26) resulted in a new omics field, i.e. peptidomics or peptidome research. Peptidomics investigations have been conducted for a number of different biological samples, including plasma (27), cerebrospinal fluid (28), saliva (29), tears (30), and brain tissues (31). However, large-scale mass spectrometry based peptidomics analysis of wound fluids has not yet been reported. As the low molecular weight peptidome could act as downstream reporters of the proteolytic action of endogenous and exogenous proteases they could serve as a promising tool for diagnosis and/or prognosis of wound healing. In a recent study, we showed that peptides from a selected protein, human thrombin, are detected and could be attributed to proteolytic actions (32). Specific thrombin-derived peptide sequences were identified in wound fluids from acute and non-healing ulcers, respectively. The result, although focusing on one single protein, demonstrated a proof-of-concept pointing at the possibility of defining peptide biomarkers for improved diagnosis of wound healing and infection. In the present study, we aimed at developing a robust peptidomics method for the characterisation of the peptidome of wound fluids. Using this method, we here compare acute non-infected wound fluids with plasma samples and find significantly higher protein and peptide numbers in wound fluids compared with plasma, which typically were also smaller in size as compared to plasma-derived peptides. Finally, we analyse wound fluids collected from dressings after facial surgery and compare three uninfected and normally healing surgical wounds with three inflamed and *S. aureus* infected wounds. We further demonstrate the utility of peptidomics in wound fluid analysis, showing peptide profiles of various selected proteins. Together, the work defines a workflow for analysis of peptides derived from human wound fluids and demonstrate a proof-of-concept that such wound fluids can be used for analysis of subtle qualitative differences in peptide patterns derived from individual patient samples.

## Results

### Comparison of sample preparation methods

To determine the most optimal method for extraction of peptides from wound fluids, wound fluids were mixed with 6 M (final concentration) urea in the absence or presence of 0.05% *Rapi*Gest or 0.1% TFA followed by 30 kDa filtration as indicated in Figure 1A. SDS-PAGE analysis of the filtrate showed high molecular weight proteins before filtration (BF) and low molecular weight peptides after filtration (Figure 1B). Urea alone (U) did only result in a few peptide bands, whereas both *Rapi*Gest (U+R) and TFA (U+T) resulted in more abundant peptide patterns. To further analyse and optimise peptide extraction, various volumes of wound fluids were filtrated as described above, followed by desalting and concentrating of defrosted samples using StageTips and finally LC-MS/MS analysis of peptides between 700 and 6400 dalton (depicted in Figure 1A) was performed. The results showed a sample volume dependent increase in the number of identified peptides and corresponding proteins for all used buffers (Figure 1C). In agreement with the SDS-PAGE results, urea alone resulted in lower numbers of identified proteins and unique peptides than the other two buffers. Interestingly, similar numbers of proteins were identified when using urea and TFA or urea and *Rapi*Gest (180 versus 173) for 100 µL of WF, whereas more unique peptides were found using the latter buffer (2931 versus 3543). Based on these results, 100 µl of wound fluids in 6M urea supplemented with 0.05% *Rapi*Gest was selected for further studies.

**Figure 1.**
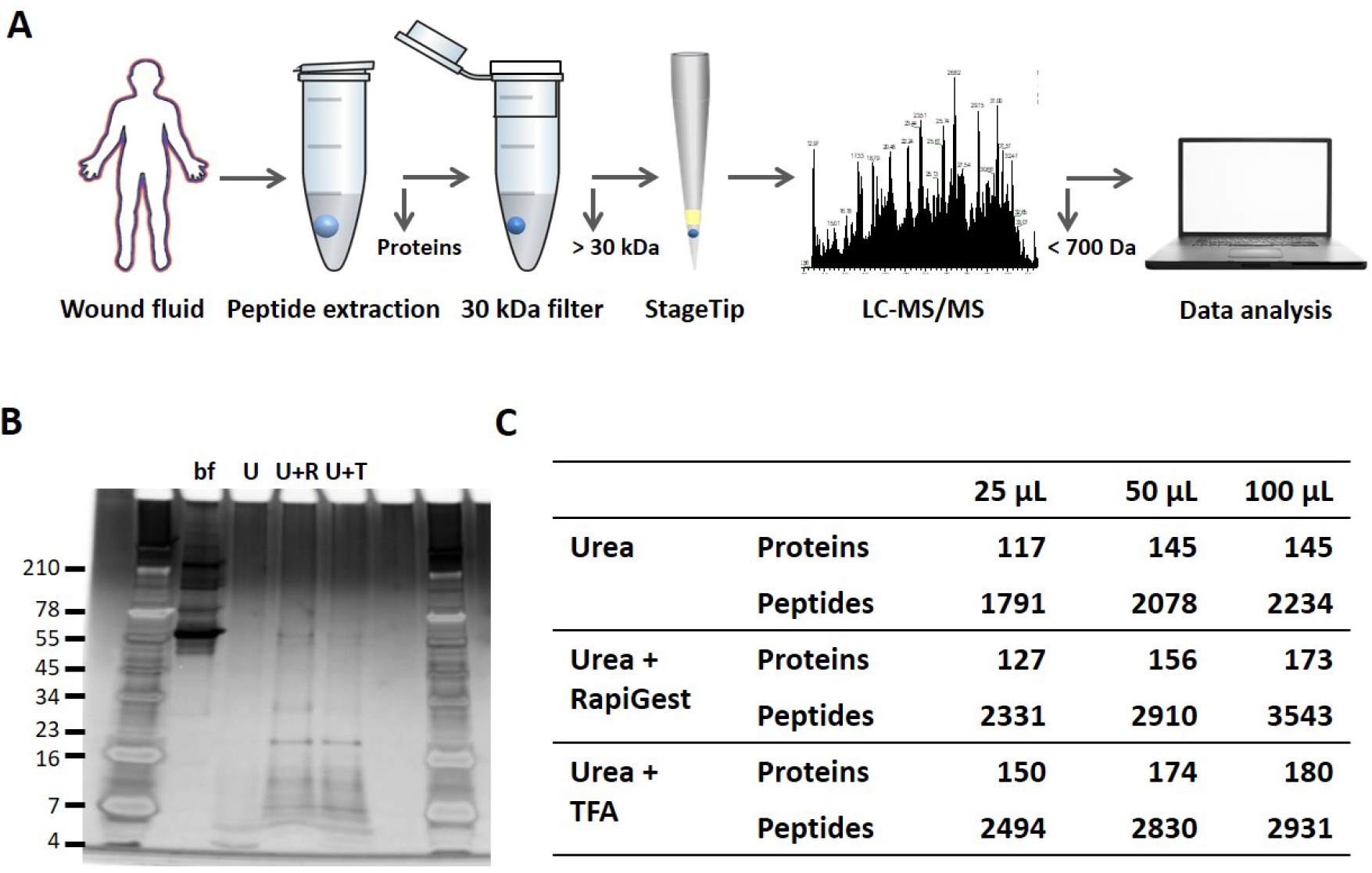
Comparison of sample preparation methods. (**A**) Schematic overview of the workflow. Peptides were extracted from 25 µl, 50 µl or 100 µl of wound fluid in 6M urea (U), 6M urea + 0.05% *Rapi*Gest (U+R) or 6M urea + 0.1% TFA (U+T), using 30 kDa cut-off filters. Stored filtrates were defrosted, followed by peptide concentration using StageTips and finally 1.6 µL of the original wound fluids were analysed by nanoLC-MS/MS. (**B**) Representative example of a 10-20% Tricine gel run with 1 µl of sample before filtration (BF) or 22.5 µl of sample after filtration, extracted from 100 µl of wound fluid, ran under non-reducing conditions and stained with SilverQuest stain. (**C**) Total numbers of identified peptides and corresponding proteins for the different buffers and amounts of wound fluid as analysed by MS. Results are shown as combined data of 2 injections per sample.

### Robustness of sample preparation method

To determine the robustness of the selected sample preparation method, each step of the workflow was investigated. The results showed no difference in peptide patterns on SDS-PAGE after dividing one sample preparation over two 30 kDa cut-off filters (Figure 2A). Nevertheless, LC-MS/MS analyses of two injections per sample showed that 62% (170) of the peptides came from the same proteins (Venn diagrams Figure 2A), whereas 58% (3363) of all unique peptides were found in both samples, indicating that the filters do interfere with peptide recovery. Next, we investigated the reproducibility of sample injection using one preparation injected three times on the same day. As shown in figure 2B, we found similar heatmap patterns for the protein scores for the three injections. Furthermore, similar patterns were also found for the number of unique peptides identified and the percentage coverage of the protein by these peptides. Moreover, comparison of one preparation injected twice on two different days resulted in clear correlations in protein score, number of unique peptides and protein coverage (Figure 2C), indicating that sample storage at 4 °C does not significantly influence the results. Finally, we tested the reproducibility of the entire sample preparation method by comparing 2 independently generated samples of the same wound fluid and found a good correlation for all three measured parameters (Figure 2D). Taken together, the above results show that the selected sample preparation method and storage is robust and can be used for peptidome comparison of various donor fluids.

**Figure 2.**
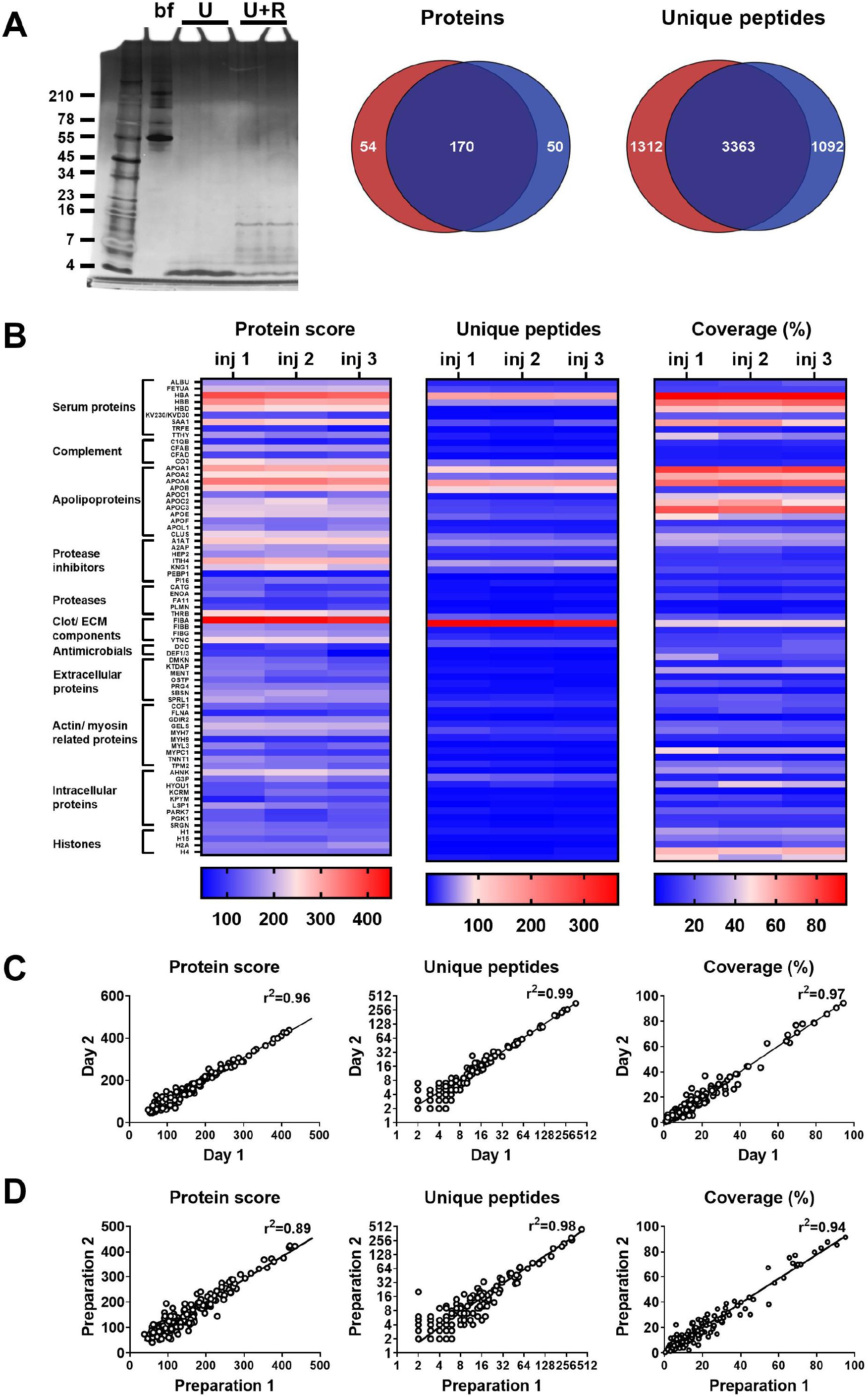
Robustness of sample preparation method. Peptides were extracted from 100 µl wound fluid in 6M urea supplemented with 0.05% *Rapi*Gest using the workflow shown in figure 1. (**A**) To investigate possible influence of the used filters on the obtained results, one wound fluid preparation was divided over two 30 kDa cut-off filters, centrifuged and filtrates were analysed using SDS-PAGE (U+R) and nanoLC-MS/MS. Combined results from 2 injections per sample are shown in Venn diagrams depicting proteins and unique peptides. For SDS-PAGE, 1 µl of sample before filtration (BF) or 22.5 µl of sample after filtration were run on a 10-20% Tricine gel under non-reducing conditions and stained with SilverQuest stain; extractions using 6M urea (U) without *Rapi*Gest are shown for comparison. (**B**) Reproducibility of sample injection using one preparation injected three times on the same day or (**C**) injected twice on two different days. (**D**) Reproducibility of sample preparation using 2 independently generated samples of the same wound fluid on different days. Combined results of 2 injections per sample are expressed as the protein score, the number of unique peptides per protein and the percentage of total coverage of each protein by the identified peptides; r-squared values are indicated in each graph.

### Comparison of plasma and wound fluids

Whereas proteases are activated and/or released in the wound environment during inflammation, plasma from healthy donors should not contain activated proteases and therefore far less peptides. Indeed, a clear difference between three wound fluids and three plasma samples could be observed on SDS-PAGE, the latter samples containing fewer distinct bands and of a higher molecular weight (Figure 3A). In agreement, LC-MS/MS analysis showed substantially more peptides in wound fluids as compared to plasma (Figure 3B). When pooling the results of the three individuals for each fluid type, over 5 times as many proteins and 6.8 times as many peptides were observed in wound fluids as compared to plasma samples (Figure 3C). Moreover, only 9.6% (30) of all proteins and 2% (121) of all peptides were detected in both types of fluids, although not necessarily in all 3 fluids in each group. Interestingly, we found relatively more small peptides (700-1500 Da) in wound fluids, whereas peptides larger than 1700 Da were relatively more prevalent in plasma (Figure 3D, right panel). Finally, heatmaps of the proteins that were either common for the three plasma samples and/or the three wound fluid samples were generated, ordered on function or localisation, showing clear differences between these two types of samples in the protein score, number of unique peptides and coverage (Figure 3E). As expected, no peptides derived from proteases were detected in plasma. Besides the number of identified peptides, another difference between plasma and wound fluids is the degree and type of post translational modifications (Supplementary Table 1). For each wound fluid, we found a higher degree of peptide modifications than in plasma. In plasma, the average deamidation of the peptides was 3.1% as compared to 6.4% for WF sample, whereas the degree of oxidation of methionine was 21.7% in wound fluids while only 4.5% in CP samples. All peptides identified in the plasma and wound fluid samples are listed in supplementary datasets 1 and 2.

**Figure 3.**
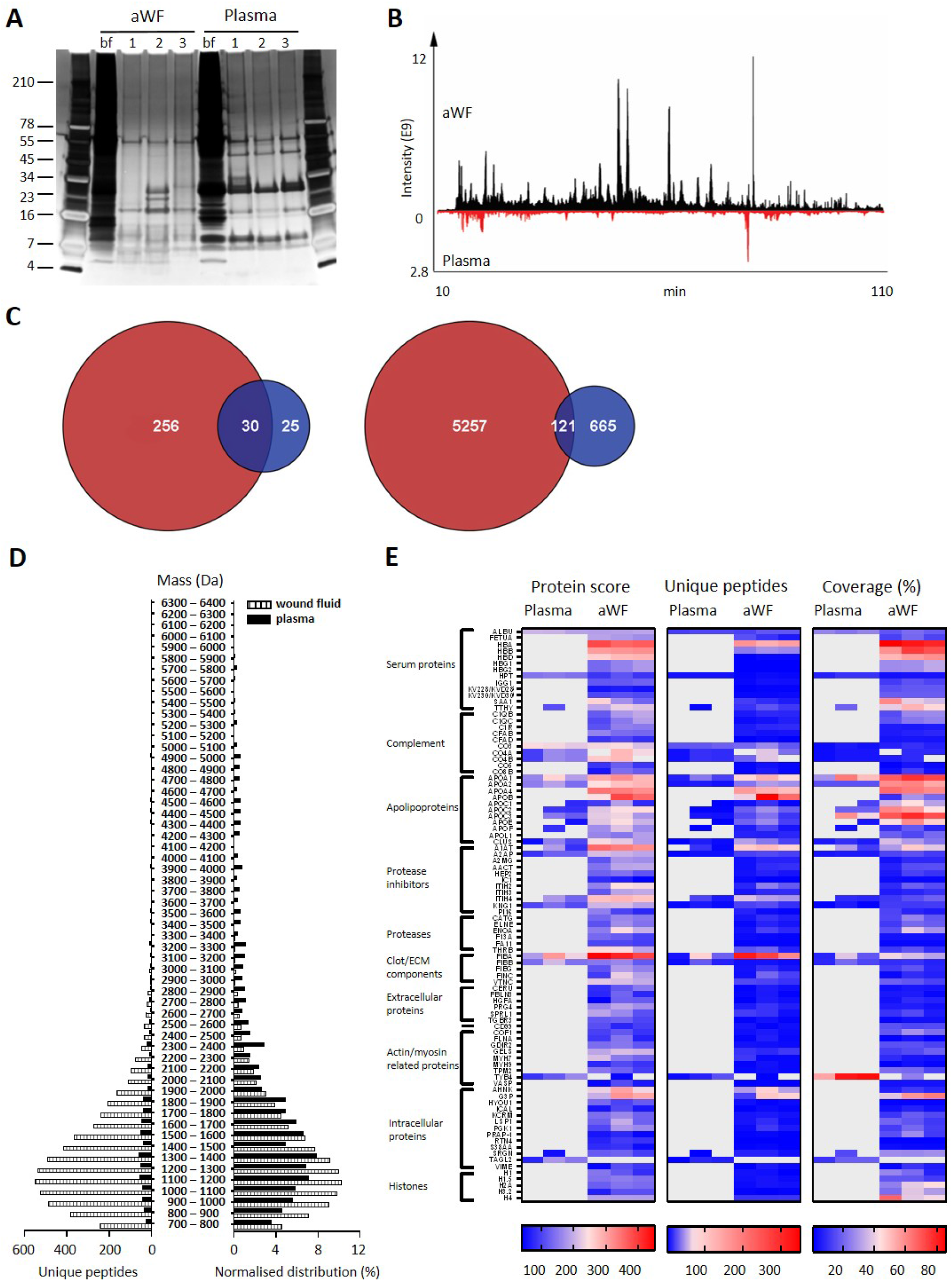
Comparison of plasma and wound fluids. Peptides were extracted from 100 µl acute wound fluid (aWF) or citrated plasma in 6M urea supplemented with 0.05% *Rapi*Gest using the workflow shown in figure 1. (**A**) Comparison of three wound fluids and three plasma samples as analysed using a 10-20% Tricine gel ran under non-reducing conditions and stained with SilverQuest stain. (**B**) Representative LC-MS/MS chromatograms of wound fluid (top) and plasma (bottom) preparations. (**C**) Comparison of the pooled results of three wound fluids with three plasma preparations using Venn diagrams depicting total number of identified proteins and unique peptides. (**D**) Distribution of peptides from representative wound fluid and plasma preparations based on molecular weight. The results are shown as total numbers (left) and normalised values (right). (**E**) Heatmaps comparing wound fluids and plasma depicting the protein score, the number of unique peptides per protein and the percentage of total coverage of each protein by the identified peptides. Results are shown as combined data of 2 injections per sample.

### Comparison of acute wound fluids

To investigate similarities of sterile acute wound fluids, five wound fluids from different donors were processed and analysed with LC-MS/MS. The data of four injections per sample (two injections a day at two different days) were merged and then subjected to a database search. For all wound fluids, high numbers of peptides were detected (ranging from 2649 to 4271, Supplementary Table 2) and these peptides corresponds to a total of 329 proteins. As illustrated in Fig 4A, 74 proteins were detected in all five wound fluid samples. Within this group of 74 proteins, there were 783 identical peptides (Fig 4A, second venn diagram), which amounts to 15% of the total peptides for the common proteins. Notably, the slightly higher number of common peptides in the venn diagram for all detected proteins (Fig 4A, third venn diagram) as compared to that found for the common proteins can be explained by the presence of 14 peptides that were not unique for one of the 74 common proteins, but were also assigned to other proteins that were not included in the common group of proteins. Interestingly, we found a difference in the average peptide length, ranging from 12.3 to 13.7 aa, of the identified peptides from the five different wound fluids (Supplementary Table 2), suggesting that the samples have been exposed to different levels of proteolytic activity. Finally, heatmaps of the proteins that were common for all five samples were generated showing resemblances in protein score, numbers of unique peptides and protein coverage (Figure 4B), which further proves the usefulness of peptidomics in wound fluid analysis.

**Figure 4.**
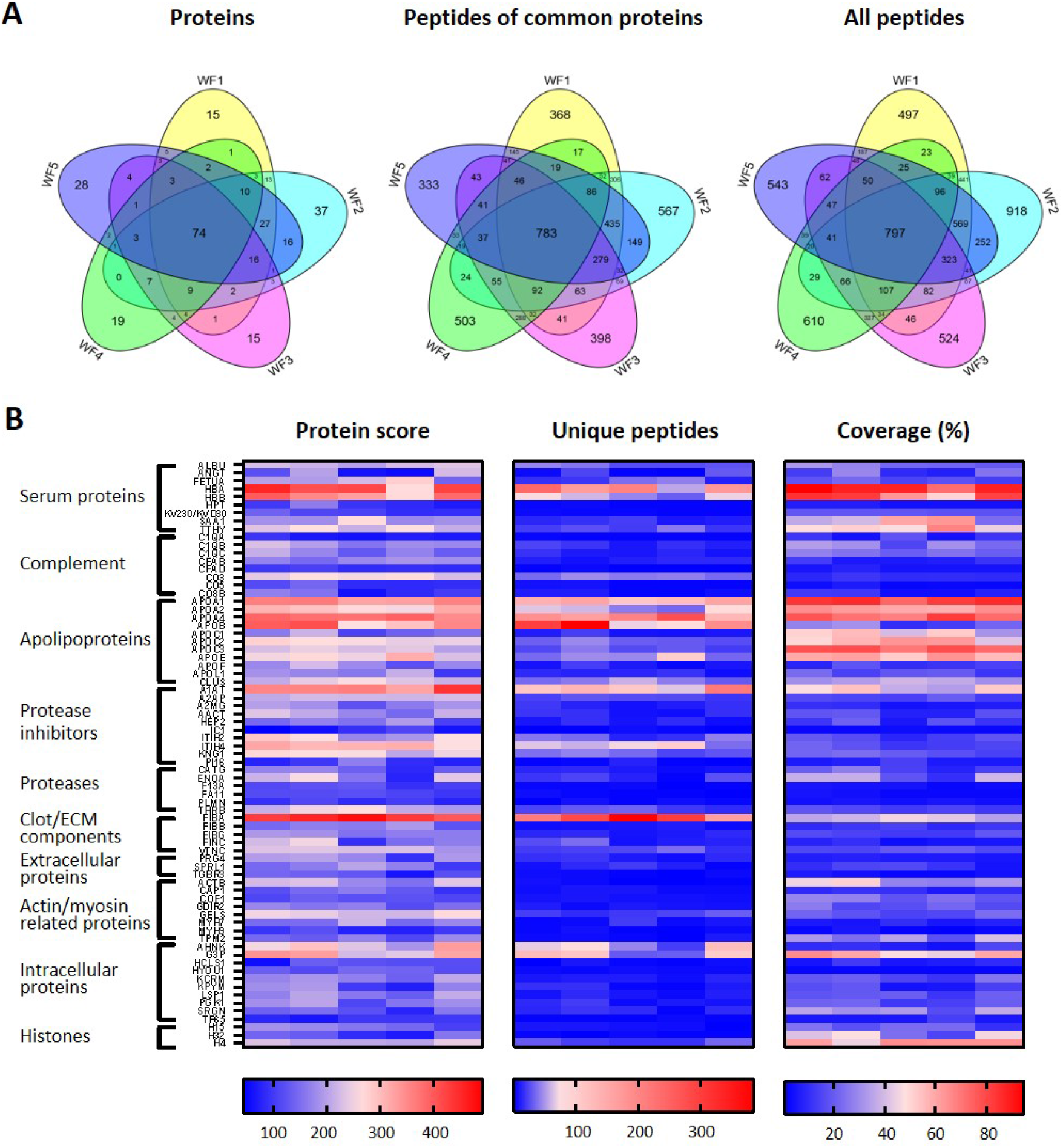
Comparison of five acute wound fluids. Peptides were extracted from 100 µl wound fluid in 6M urea supplemented with 0.05% *Rapi*Gest using the workflow shown in figure 1. (**A**) Comparison of five wound fluids using Venn diagrams depicting proteins, unique peptides of the 74 proteins common for all five wound fluids and unique peptides of all identified proteins. (**B**) Heatmaps comparing the five wound fluids depicting the protein score, the number of unique peptides per protein and the percentage of total coverage of each protein by the identified peptides. Results are shown as combined data of 4 injections per sample.

### Comparison of non-inflamed, non-infected wounds with inflamed and infected wounds

Finally, and as a proof-of-concept, we selected 6 patients from a previously published clinical study (15), who had undergone facial full-thickness skin grafting, and either had healed well at the 7 day follow-up after surgery (Figure 5A, low inflammation group) or had an inflamed wound infected by amongst others *Staphylococcus aureus* (high inflammation group). Cytokine analysis of extracts made of the wound dressings indeed showed increased levels of IL-1β, IL-6, IL-8 and TNF-α in the high inflammation as compared to the low inflammation group (Figure 5B). Further analyses of the dressing extracts using SDS-PAGE (Figure 5C) and zymograms (Figure 5D) showed a positive correlation between protein degradation and enzymatic activity, both more apparent in the high inflammation group. Next, two independently generated sample preparations were made of each extract, using the sample preparation method above, and subjected to LC-MS/MS. The data of four injections per sample (2 injections for each of the duplicate extracts) were merged and then subjected to a database search. All identified peptides are listed in supplementary dataset 3. Interestingly, when pooling the results of the three patients per group, we found that 163 proteins (2666 peptides) were found in both groups, although not necessarily in each single patient, whereas 156 proteins (5273 peptides) were only found in the low inflammation group versus 90 (4661 peptides) in the high inflammation group (Figure 5E). These results suggest a higher degree of proteolysis in the high inflammation group, as per identified protein an average of 52 unique peptides was found as compared to 34 in the low inflammation group. Furthermore, the individual samples within the high inflammation group show a higher degree of variability as only 23% of the proteins (10% of all unique peptides) were found in all three patient dressing extracts versus 34% (16% of the peptides) for the low inflammation group (Supplementary Figure 1). This variability is further visualised in heatmaps, generated of the proteins that were found in all three extracts of either the low inflammation group or the high inflammation group (Figure 5F). As expected, some proteins, such as the serum proteins albumin (ALBU) and hemoglobin (HBA/HBB/HBD), are identified in all wound fluids and dressing extracts (heatmaps Figure 2-5), whereas complement component C9 (CO9), which is a constituent of the membrane attack complex that plays a key role in the innate immune response against bacteria, is only found in fluids from infected wounds. Interestingly, the antimicrobial protein dermcidin (DCD), constitutively expressed by eccrine sweat glands, was detected in the low inflammation group, whereas the protein was absent in the high inflammation group. This may be explained by the reported proteolytic degradation of dermcidin by staphylococci (33), to such an extent that the remaining peptides can no longer be detected by LC-MS/MS. Contrastingly, the antimicrobial protein lactoferrin (TRFL) was only detected in the high inflammation group. As lactoferrin is secreted by neutrophils upon degranulation, and influx of neutrophils is extensive in inflamed wounds, these results are as expected.

**Figure 5.**
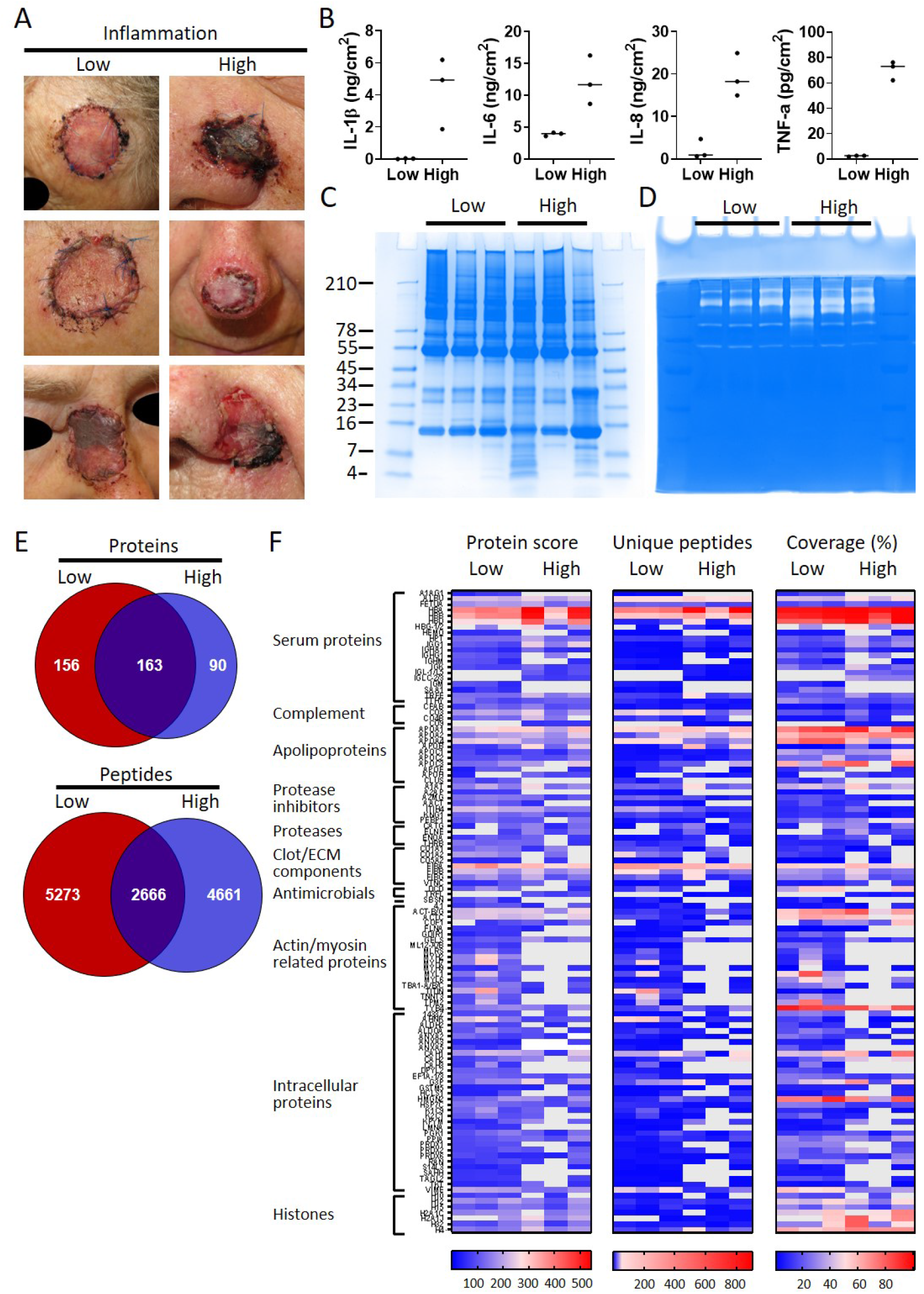
Comparison of healing and non-healing infected wounds. (**A**) Photos of wounds, seven days after surgery, of six patients who had undergone facial full-thickness skin grafting. On the left side are the three that healed well and showed low inflammation and no infection, while on the right side wounds are depicted that were highly inflamed and infected with a.o. *Staphylococcus* aureus. Dressing extracts were made of the seven day old dressings derived from each wound and analysed for cytokine content (**B**), protein and peptide composition using SDS-PAGE (**C**), and enzymatic activity using zymograms (**D**). Peptides were extracted from 280 µg of wound dressing extract in 6M urea supplemented with 0.05% *Rapi*Gest using the workflow shown in figure 1. (**E**) Comparison of the pooled results of the three low inflammation samples with the three high inflammation samples using Venn diagrams depicting total number of identified proteins and their unique peptides. (**F**) Heatmaps comparing the six samples depicting the protein score, the number of unique peptides per protein and the percentage of total coverage of each protein by the identified peptides. Results are shown as combined data of 2 independent sample preparations with 2 injections per sample.

To further investigate protein fragmentation, peptide profiles and peptide alignment maps were generated in order to visualize the qualitative changes at the peptide level. Figure 6-8 show selected peptide profiles representing proteins of relevance for fibrin formation, wound healing and inflammation, as well as antimicrobial defence. Notably, these profiles do not contain peptides that are post translationally modified. Furthermore, the depicted peptide alignment maps are only showing a selection of the sequences detected for the protein areas that are highlighted by blue boxes.

**Figure 6.**
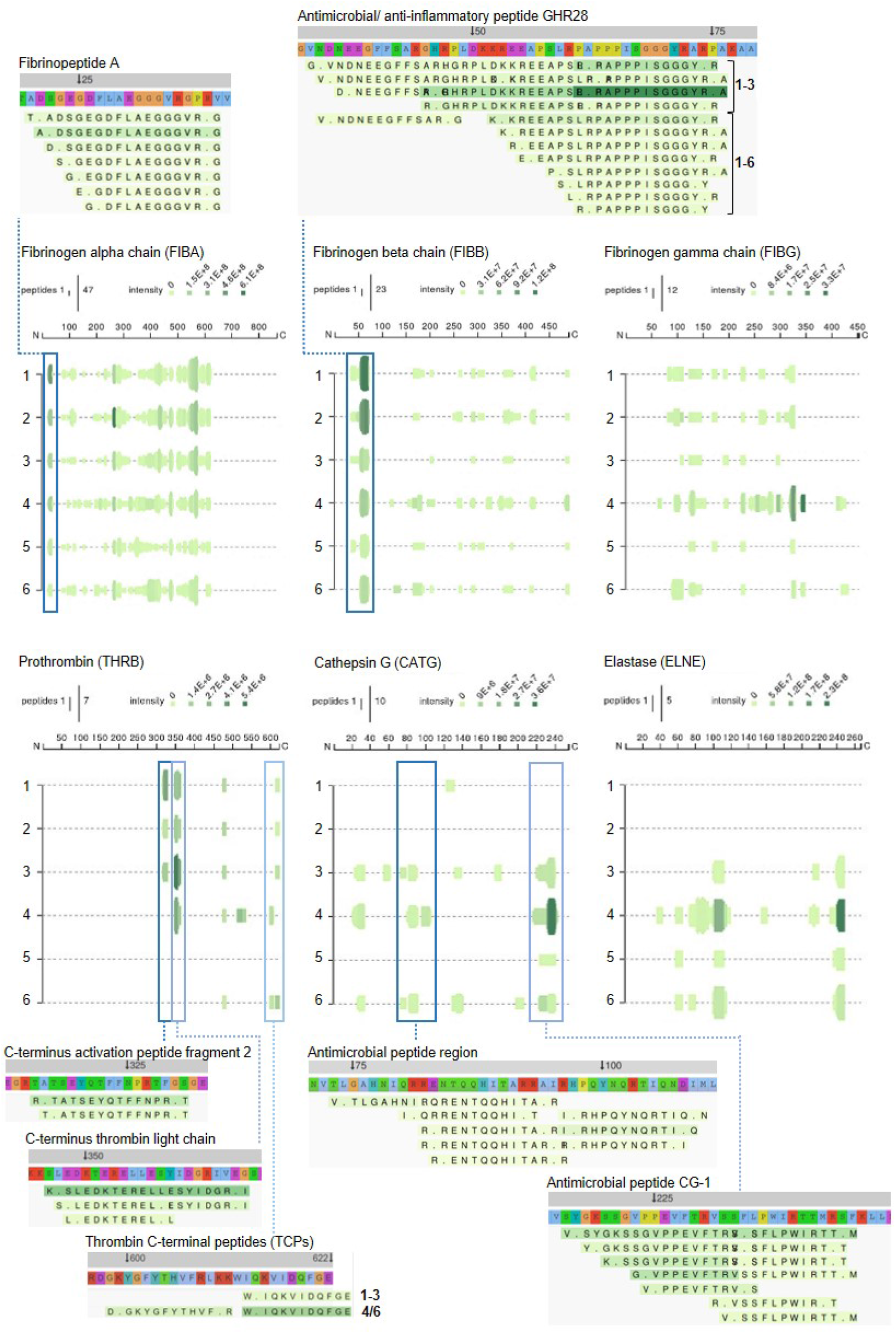
Peptide profiles of fibrinogen and selected proteases Peptide profiles and peptide alignment maps of three fibrinogen chains and the proteases prothrombin, cathepsin G and neutrophil elastase, were generated from the UniProt IDs, peptide sequences, start and end, and intensities for each protein using the web application Peptigram. The height of the green bars is proportional to the number of peptides overlapping the amino acid residue, while the intensity of the color (green) is proportional to the sum of the intensities overlapping this position. Interesting peptide regions are highlighted by blue boxes, and of the corresponding peptides one sequence of each identified N-terminal is shown for illustration purposes; 1-3, low inflammation samples, 4-6, high inflammation samples.

As expected, peptides derived from the three different fibrinogen chains alpha, beta, and gamma were identified (Figure 6). Peptides similar to clinically relevant fibrin degradation products, composed of fibrinopeptides, were found. Furthermore, the antimicrobial and anti-inflammatory peptide GHR28 from the beta chain (34) was identified in the three non-inflamed wounds, whereas only smaller fragments were detected in the inflamed and infected wounds. Prothrombin is a major protease activated during wound healing, and it was notable that peptides from the thrombin light chain were observed in all three wound samples from non-infected normally healing wounds, whereas they were absent in two of the infected wound fluids. Furthermore, fragments derived from thrombin-derived C-terminal peptides (TCP), previously found to exert antimicrobial and immunomodulatory effects in wounds (32, 35-37), were identified. Whereas thrombin is a major liver-derived enzyme, others such as cathepsin G and elastase are secreted from neutrophils during inflammation and infection. In agreement, peptide fragments from these two enzymes were particularly observed in samples from infected wounds. For cathepsin G, peptides were identified from two antimicrobial regions (38), the latter consisting of C-terminal fragments corresponding to the previously described antimicrobial peptide CG-1 (39). Interestingly, a similar C-terminal fragmentation pattern was identified in neutrophil elastase. Taken together, these results indicate that specific degradation patterns of major plasma and neutrophil proteases can be detected using wound extracts derived from dressings after surgery.

High molecular weight kininogen (HMWK) is a multifunctional 120-kDa glycoprotein found in plasma and in granules of platelets (40). The protein is composed of six domains, each having different properties and specific ligands (41). Whereas domains 2 and 3 have cysteine protease inhibitor activities, the D4 domain contains the bradykinin sequence, which is released by plasma kallikreins during contact activation and actions of proteases such as neutrophil elastase (42). Thus, limited proteolysis of HMWK generates highly vasoactive and pro-inflammatory peptides, which are formed at sites of tissue injury and inflammation. The cell-binding D5 from HMWK contains regions dominated by histidine, glycine, and in certain parts, interspersed lysine residues, which also can mediate antimicrobial effects (43). As seen in Figure 7, these peptide fragments were indeed observed in the postoperative wounds. Further evidence for the usefulness of the methodology is provided by the detection of fragmentation patterns from additional protease inhibitors such as alpha-1-antitrypsin, antiplasmin, as well as inter-alpha-inhibitor, all yielding peptide sequences from distinct regions of these inhibitors.

**Figure 7.**
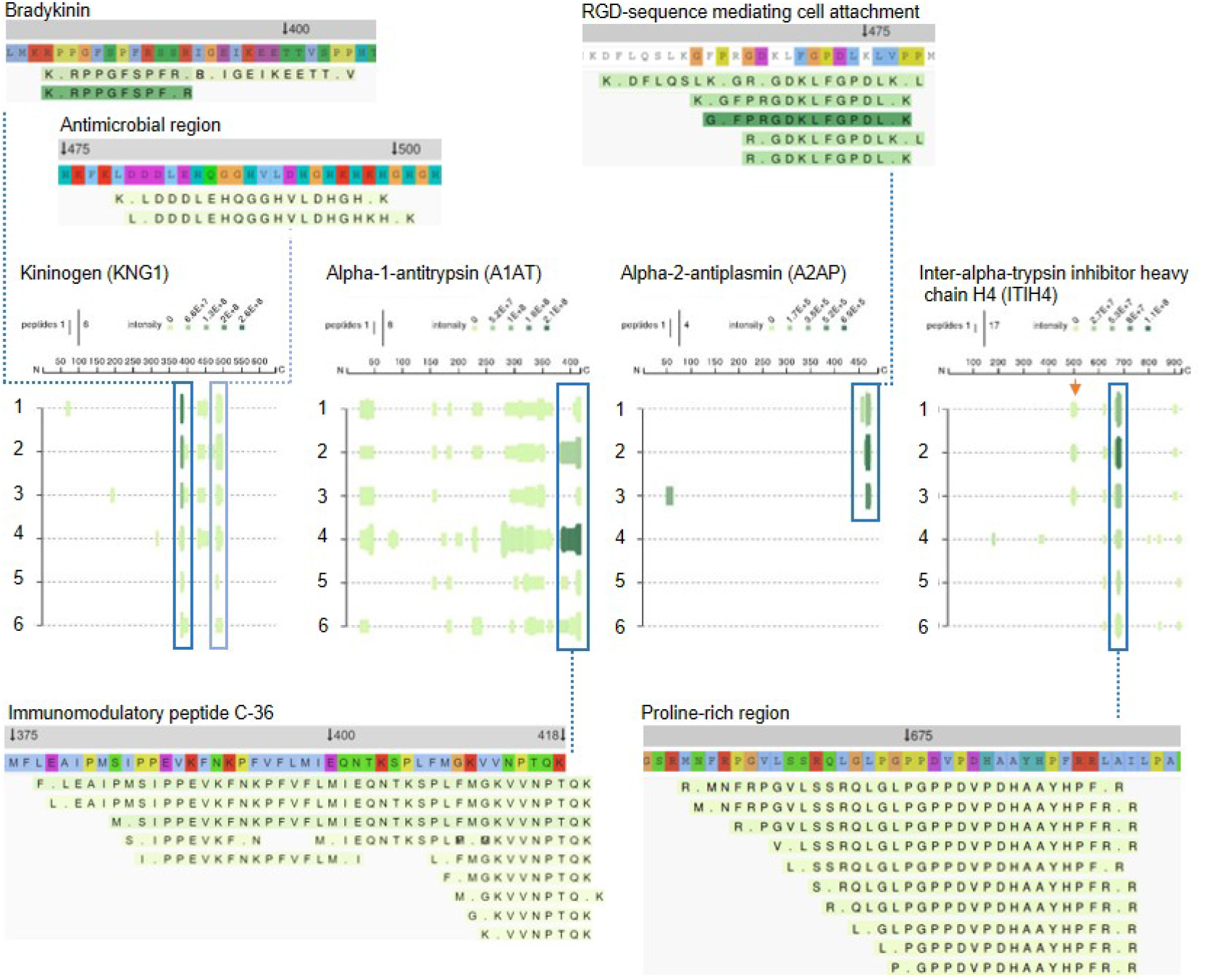
Peptide profiles of selected protease inhibitors Peptide profiles and peptide alignment maps were generated for the protease inhibitors kininogen, alpha-1-antitrypsin, alpha-2-antiplasmin and inter-alpha-trypsin inhibitor heavy chain H4. Interesting peptide regions are highlighted by blue boxes, and a selection of the corresponding peptides is shown for illustration purposes; 1-3, low inflammation samples, 4-6, high inflammation samples. The orange arrow indicates non-highlighted peptide sequences unique for the three low inflammation samples.

Cleavage of A1AT occurs at the reactive center loop (RCL) by proteases such as neutrophil elastase, although MMPs and *S. aureus* enzymes can also cleave the antiproteinase (44, 45). Hence A1AT may be reporting wound derived protease activity. Moreover, cleavage between Phe^376^-Leu^377^ generates a carboxy-terminal 42-residue peptide, and variants of this fragment, such as a C-terminal 36 aa peptide have been identified in several human tissues, where they may exert various immunomodulatory functions (46, 47). Notably, such fragments were also identified in the wound fluids from surgical wounds. Interestingly, C-terminal regions of antiplasmin were only observed in normally healing wounds, and it is notable that this region of plasmin contains bioactive epitopes that can modulate urokinase activity (48) and interact with endothelial cells (49). Recently, C-terminal fragments generated *in vivo* were indeed identified (50). Inter-alpha-trypsin inhibitor heavy chain H4 (ITIH4) is a 120 kDa serum glycoprotein secreted primarily by the liver. Peptides derived from the proline-rich region may be biomarkers for a variety of disease states including breast cancer (51), further illustrating that surgical wounds contain regions of biological and diagnostic importance. Notably, peptides from residues 488-504 were only identified in normally healing wounds (Figure 7, orange arrow).

In addition to the proteases and protease inhibitors above, the complement cascade is another fundamental defense system activated during wounding and infection. Factor B is part of the alternate pathway of the complement system is cleaved by factor D into 2 fragments: Ba and Bb. Bb, a serine protease, then combines with complement factor 3b to generate the C3 or C5 convertase. Inspection of the peptide profile for this factor showed that fragments from particularly the S1 peptidase domain were present in the infected wound (Figure 8). For the major complement component 3 (CO3), a series of similar fragmentation patterns were identified, with peptides originating in different regions of CO3. Importantly, N-terminals of the well-known anaphylatoxin C3a which harbors antimicrobial activity (52), as well as the opsonin C3b were identified. Besides common fragments found for all 6 patients, 2 peptide sequences were only found in normally healing wounds (orange arrows). As mentioned, complement component 9, which is part of the membrane attack complex that plays a key role in the immune response against bacteria, was only detected in fluids from infected wounds.

**Figure 8.**
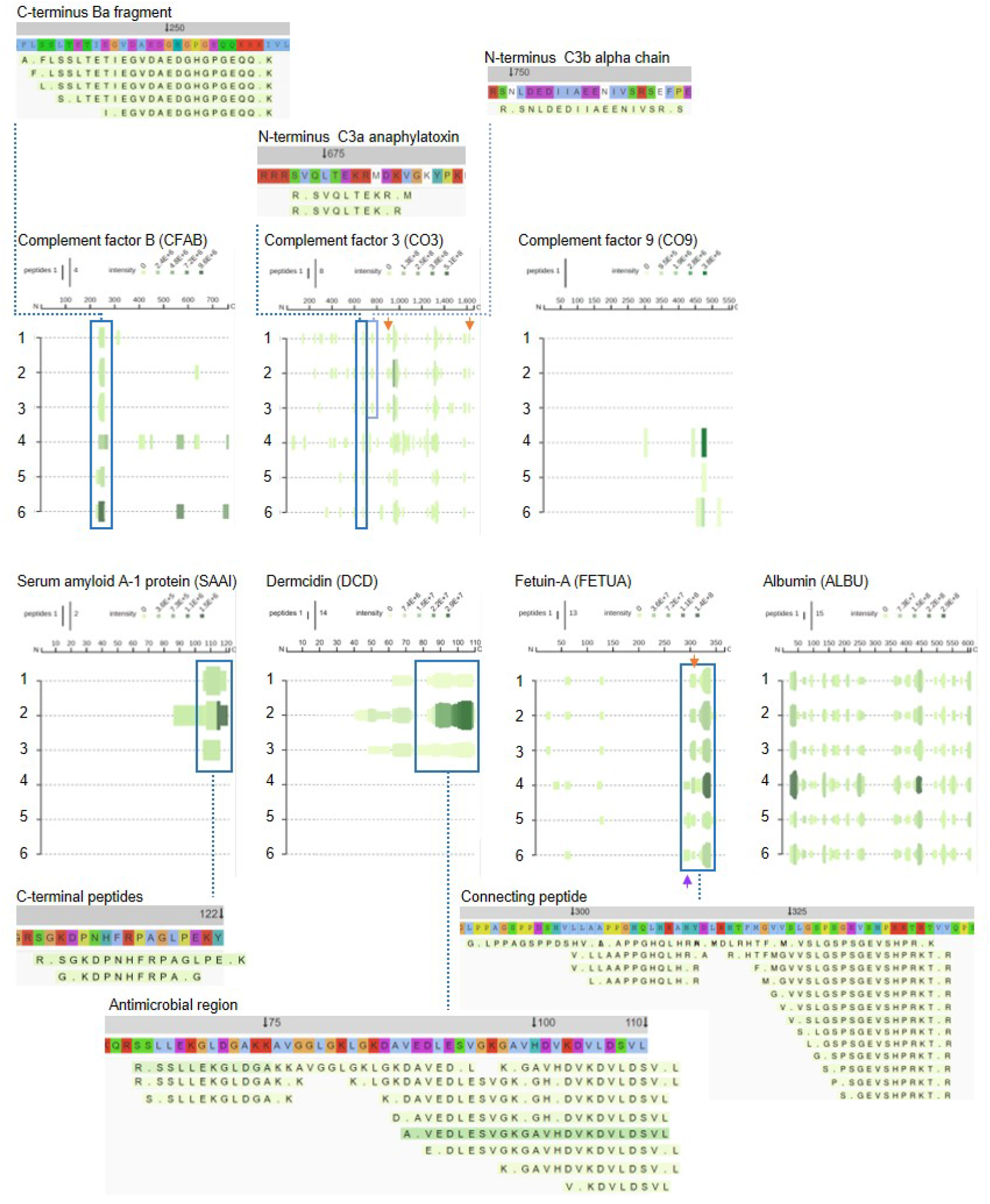
Peptide profiles of selected complement factors and additional proteins. Peptide profiles and peptide alignment maps were generated for the complement factors B, C3 and C9, as well as the proteins serum amyloid A-1, dermcidin, fetuin-A and albumin. Interesting peptide regions are highlighted by blue boxes, and a selection of the corresponding peptides is shown for illustration purposes; 1-3, low inflammation samples, 4-6, high inflammation samples. The orange arrows indicate non-highlighted peptide sequences unique for the three low inflammation samples, while the purple arrow indicates those of the high inflammation group.

Further evidence for distinct proteolytic events affecting selected proteins is seen in the peptigrams representing serum amyloid A-1 (SAA1) peptides, as well as dermcidin (DCD)-derived fragments, which both were only found in non-infected wounds (Figure 8, lower panels). SAA1 can upregulate inflammatory cytokines, whereas SAA1 peptides suppress inflammation (53). Moreover, the C-terminal SAA(86-104) binds and inhibits human cystatin and carboxyterminal fragments are involved in amyloid diseases (54). Interestingly, similar C-terminal fragments were found in non-infected wounds, indicating that SAA is proteolyzed in wounds, suggesting a yet undisclosed physiological role for such fragments during normal wound healing. DCD is an antimicrobial peptide found in skin (55). Proteolytic processing of DCD gives rise to several truncated peptides (56). Interestingly, variants of the peptide DCD-1L having the N-terminal SSLLEK (57) were indeed identified in normally healing surgical wounds. A proteolytically processed form of fetuin-A, also denoted α2-HS glycoprotein, lacking a segment of 40 amino acid residues bridging its heavy and light chain portions (“connecting peptide”) has been described, suggesting that this peptide is released by post-translational processing to fulfil biological role(s) of fetuin-A. The connecting peptide region, which is highly susceptible to proteolytic breakdown *in vitro* and *in vivo* (58), was present in the surgical wound fluids. Interestingly, peptides starting with the N-terminal LPPAGS were only detected in the three inflamed wounds (purple arrow), whereas those starting with LLAAPP were unique for the three non-inflamed wounds (orange arrow). Finally, to show the validity and robustness of the method, the peptide profile of serum albumin is presented, showing a high similarity between the fragmentation patterns in the individual wounds.

## Discussion

Endogenous peptides serve as biomarkers of disease progression for several different diseases, and the here presented strategy and methodology identified the wound fluid peptidome as a source for assessment of wound fluid dynamics, as these fragments represent proteolytic events that are the result of basic physiological processes involving coagulation and complement activation, but also additional proteolytic activities by endogenous and bacterial proteases. Degradation of proteins occurs due to the action of set of a proteases and biological processes that results in a low molecular weight fraction of endogenous peptides with specific cleavage points. For the analysis of the wound fluid peptidome by mass spectrometry it is necessary to extract and enrich the peptides, thereby excluding proteins and other interfering molecules. Several different protocols and methods are available for extraction of peptides from biofluids prior to the mass spectrometry measurement. In our work we have chosen to investigate the peptidome of wound fluids by a combination of low molecular weight filtration, solid phase purification and enrichment followed by mass spectrometry detection and database search. Although solubility of the peptides and the binding of peptides to proteins will affect the outcome of the filtration procedure, previously it was found that solubilisation using urea was efficient for the extraction recovery of endogenous peptides (31). The LC-MS/MS analysis was performed using data-dependent analysis, where the instrument is set to sequence as many peptides as possible for each sample and the same sample was injected in duplicates. The chosen method allowed us to characterise more than 7700 peptides in acute wound fluids derived from the degradation of over 370 proteins. We have focused our subsequent analysis on a subset of peptides corresponding from the degradation of the most abundant proteins that could be measured in all of the examined acute wound fluids. The definition of the wound peptidome of acute wounds in well-defined sterile wound fluids validated the approach, identifying multiple peptides derived from several protein families. The following analyses on dressing extracts from skin grafted surgical wounds, provided a proof-of-principle showing the possibility of analysis of selected peptide regions as potential biomarkers for inflammation and wound healing. This is of high relevance from a clinical perspective, as skin grafting surgery is normally associated with a higher risk of SSI (3, 15) motivating the use of such wound fluids from this type of dermatologic surgery.

As briefly mentioned in the introduction, classical proteomics approaches have been employed in the study of different wound types. Eming et al. studied the distribution of tissue repair proteins in exudates of healing acute and non-healing venous wounds on the legs (11). Subsequent studies have added to the categorization of proteins identified in various wounds such as diabetic and pressure ulcers (59-63). Auf dem Keller and colleagues applied quantitative proteomics strategies to dissect proteolytic pathways during wounding and the activity of distinct protease groups along the healing process. Moreover, they also mapped proteolytic pathways *in vivo* and established protease-substrate relations that will help to better understand protease action in cutaneous wound repair and other inflammatory conditions in skin (22, 23, 64, 65). However, it is of note that methods for detection of down-stream products reflecting inflammation are rare. Although Auf dem Keller’s work involves studies on such protease patterns, their method focuses on “N-terminomics”, while low molecular weight peptides, such as the ones studied here, are mainly lost during the preparation steps. Interestingly, at the proteomic level, in their studies on healing and non-healing wounds on the legs, Eming et al showed that the serine protease thrombin as well as the antimicrobial DCD was exclusively detected in healing wounds (11). In contrast, the neutrophil derived lactoferrin was increased in non-healing wounds. Moreover, the three major fibrinogen chains alpha, beta, and gamma, were dominating in the wound fluids from all wound types. Although the wounds types are obviously different with respect to localisation and pathogenesis, the clinical grading into non-healing and healing is the same for both studies. It is therefore notable that corresponding observations on the above proteins were indeed made in this study at the peptidome level as well, lending further support for the diagnostic potential of the here described methodology.

Today, the current method for characterization of infection and related wound complications in the clinic is first to assess wound status according to clinical experience combined with bacterial detection. There are many obvious signs of advanced infection including redness, heat, swelling, purulent exudate, smell, and pain, but the challenge is to translate these observations to objective and sensitive methods that correctly evaluate wound status and in particular, the associated inflammation, before wounds even reach this dysfunctional state. Moreover, presence of bacteria in wounds does not correlate with wound inflammation and infection. For example, non-healing ulcers are frequently colonized by staphylococci, even without infection, and postsurgical wounds can also harbour *S. aureus* without signs of clinical infection (15, 66, 67). Therefore, given that mere bacterial presence is not discriminatory for wound infection, there is a clear need for objective methods that enable detection of dysfunctional wound healing to better guide the use of treatment. Approaches under development today to evaluate bacteria and the concomitant inflammation are use of biological or chemical sensors of wound exudates to detect bacterial antigens, monitor pH, temperature, oxygen and enzymes. Spectroscopic and imaging techniques are also possible as future advanced wound monitoring techniques (68-71). Methods that measure host responses as one way to assess wound healing, and particularly inflammation, have also been developed. For example, the major enzymes from neutrophils, human neutrophil elastase (HNE) and cathepsin G (CatG) have been reported as early stage warning markers for non-healing ulcers. In the clinic, detection of HNE or MMPs is used by a device developed by Systagenix, measuring elevated protease activity through their WoundcheckTM Protease Status diagnostic. In a recent study, it was identified that wounds that had a clinical appearance of being more inflamed indeed had higher levels of proteases, as demonstrated for the major MMPs, MMP-2 and −9. These wounds had increased cytokine levels relative to the wound surface area, particularly observed for IL-1ß, but also for TNF-α (15). Moreover, these wounds also had significantly higher overall bacterial loads, a factor of importance in the development of SSIs. As demonstrated here, apart from the cytokines IL-1β, IL-6, IL-8 and TNF-α, a vast number of potential new peptide sequences could be detected, hence providing a proof-of-principle for utilization of wound fluid peptidomics in future biomarker-oriented studies on larger well-defined patient groups. Combining the use of such new peptides as biomarkers along with classical analyses of cytokines and MMPs could yield higher diagnostic sensitivity and specificity with respect to wound status and infection risk. Moreover, the development of sensors, based on antibodies or aptamers that targets selected peptides could make this diagnostic approach more attractive in the clinics, enabling fast readouts and thus facilitate the needed clinical studies. Finally, the peptidomics data generated here provides previously undisclosed data on proteolytic fragmentation patterns during wounding, which may aid in the discovery of novel bioactive peptides and elucidation of their biological roles during wound healing.

## Materials & Methods

### Sample collection

Plasma was collected from citrated venous blood from healthy donors by centrifugation at 2000 x g. Sterile acute wound fluids (aWF), obtained from surgical drainages after mastectomy, were collected for 24 h, 24 to 48 h after surgery followed by centrifugation as described previously (72). Wound fluids from patients that underwent facial full-thickness skin grafting were extracted from Mepilex® (Mölnlycke Health Care, Sweden) tie-over wound dressings, which had been on the wound for one week, as described (15). In short, wound exudate was extracted from dressings after incubation in 10 mM Tris (pH 7.4) for 1 h at 8 °C while shaking. All samples were stored at −20 °C before use.

### Peptide extraction

Frozen plasma and/or sterile acute wound fluids were defrosted and mixed with 3 parts freshly made 8 M urea (in 10 mM tris, pH 7.4, yielding a final urea concentration of 6 M) alone or supplemented with *Rapi*Gest SF (0.05% final concentration; Waters, USA) or trifluoroacetic acid (0.1% final concentration; Sigma-Aldrich, Germany) and incubated for 30 min at room temperature (RT) followed by size exclusion. For this purpose, centrifugal filters with 30 kDa cut-off (Microcon 30, regenerated cellulose, Millipore, Ireland) were first rinsed with 100 µL buffer and centrifuged at 14000 g for 15 min at RT, then loaded with the incubated sample mixtures, followed by 30 min centrifugation (14000 g at RT) and a final filter washing step with 100 μL of extraction buffer (5 min, 14000 g). Due to large variations in protein content of the dressing extracts (between 2.8-16.5 mg/mL), protein concentrations were standardized. For this purpose, 10 mM Tris was added to 280 µg of each fluid to obtain a sample volume of 100 µl in total, which was then incubated with 300 µl 8 M urea supplemented with *Rapi*Gest SF for 30 min, followed by the size exclusion steps as described above. For all samples, the two filtrates containing the peptides were pooled and analysed by SDS-gel electrophoresis directly or stored at −20 °C before analysis by liquid chromatography tandem mass spectrometry (LC-MS/MS).

### SDS-gel electrophoresis

Extracted peptide samples or 20 µg of each dressing extract were denatured at 85 °C for 5 min in 1x SDS sample buffer followed by separation on 10-20% Tris-Tricine mini gels in 1x Tris-Tricine SDS running buffer for 90 min at 125V. Gels and buffers were derived from Invitrogen™ (USA). Gels were stained using GelCode™ Blue Safe Protein Stain (Thermo Scientific™, USA) or the SilverQuest™ Silver Staining Kit (Invitrogen™) according to manufacturer’s instructions and patterns were visualised using a Gel Doc™ Imager (Bio-Rad Laboratories, USA).

### Zymograms

Gels were prepared consisting of a separation gel (0.1% (w/v) gelatine, 0.1% (w/v) SDS, 10% acrylamide in 375 mM Tris buffer (pH 8.8), 0.05% (v/v) TEMED and 0.05% (w/v) APS) and a stacking gel (0.1% (w/v) SDS, 4% acrylamide in 125 mM buffer (pH 6.8), 0.1% (v/v) TEMED and 0.05% (w/v) APS). Dressing extracts (5 µg) were mixed with sample buffer (20% (v/v) glycerol, 5% (w/v) SDS, 0.03% (w/v) bromophenol blue, 0.4 M Tris-HCl pH 6.8) in a 1:1 ratio, transferred to the slots, and gels were run in electrophoresis buffer (25 mM Tris, 0.2 M glycine and 0.5% (w/v) SDS in H_2_O at pH 8.7) for 60 min at 150 V. Next, gels were washed in H_2_O, incubated for one hour in 2.5% Triton X-100, washed again and placed in enzyme buffer (5 mM CaCl_2_, 1 µM ZnCl_2_, 200 mM NaCl and 50 mM Tris-HCl pH 7.5). After overnight incubation at 37 °C while shaking (50 rpm), gels were washed and stained using Coomassie brilliant Blue. Enzymatic activity was observed after destaining the gels with a solution consisting of 10% EtOH and 14% acetic acid in H_2_O.

### Measurement of cytokine levels

Cytokine levels in the dressing extracts were determined using ELISA kits (R&D systems, USA) according to manufacturer’s instructions. Results, adjusted to the total protein concentrations of the extracts, are mean values of triplicate measurements.

### LC-MS/MS analysis

Prior to LC-MS/MS analyses, 80 µl of defrosted peptide extracts were acidified with 5 µL of 10% formic acid and then trapped and enriched on StageTip columns (73). Next, peptides were extracted with 70% ACN and 0.1% TFA and dried down before reconstitution in 20 µL of 2% ACN and 0.1% TFA. For each run, 2 µL of reconstituted sample was injected, which corresponded to 1.6 µL of the original wound fluid or plasma sample. For the evaluation of the sample preparation method, LC-MS/MS analyses were carried out on an Orbitrap Fusion Tribrid MS (Thermo Scientific) as described earlier (74), with the following modifications. During the elution steps, the percentage of solvent B increased from 5% to 22% in the first 20 min, then increased to 20% in 85 min, then to 30% in 20 min, and to 90% in a further 5 minutes, where it was kept for 5 minutes.

Subsequent analysis of patient samples was performed using an HFX Orbitrap MS system (Thermo Scientific) equipped with a Dionnex 3000 Ultimate HPLC (Thermo Fisher, USA). Injected peptides were trapped on an Acclaim PepMap C18 column (3 µm particle size, 75 µm inner diameter x 20 mm length). After trapping, gradient elution of peptides was performed on an Acclaim PepMap C18 column (100 Å 2 μm, 150 mm, 75 μm). The outlet of the analytical column was coupled directly to the mass spectrometer using a Nano Easy source. The mobile phases for LC separation were 0.1% (v/v) formic acid in LC-MS grade water (solvent A) and 0.1% (v/v) formic acid in acetonitrile (solvent B). Peptides were first loaded onto the trapping column and then eluted to the analytical column with a gradient. The percentage of solvent B increased from 2% to 27% in the first 107 min, then increased to 32% in 10 min, then to 90% in 15 min where it was kept for 5 minutes. The peptides were introduced into the mass spectrometer via a Stainless steel emitter 40 mm (Thermo Fisher) and a spray voltage of 1.9 kV was applied. The capillary temperature was set at 275 °C. Data acquisition was carried out using a top 20 based data-dependent method. MS was conducted in the range of 350–1350 *m/z* at a resolution of 120,000 FWHM. The filling time was set at a maximum of 100 ms with limitation of 3 ⨯ 10^6^ ions. MSMS was acquired with a filling time maximum 300 ms with limitation of 5⨯ 10^4^ ions, a precursor ion isolation width of 2.0 *m/z* and resolution of 15,000 FWHM. Normalized collision energy was set to 28%. Only multiply charged (2+ to 5+) precursor ions were selected for MS2. The dynamic exclusion list was set to 30 s.

### Data analysis

MS/MS spectra were searched with PEAKS software. UniProt Human, including 20413 protein sequences, was used with nonspecific cleavage; 5 ppm precursor tolerance and 0.5 Da fragment tolerance were used for the experiments conducted with the Fusion instrument and 0.02 Da for the fragments when the HFX was used. Oxidation (M) and deamidation (NQ) were treated as dynamic modification. Search result were filtered by using 1% FDR for the wound fluids, or a 23 score for the plasma samples, and at least 2 unique peptides for each protein. All data are uploaded to the proteomeXchange repository.

For data visualisation, graphs and heatmaps were generated using Graphpad Prism, venn diagrams were made in VennDis (75), while peptide profiles and peptide alignment maps were made using Peptigram software (76).

### Study approval

This study was carried out in accordance with the recommendations of the Ethics Committee at Lund University, Lund, Sweden with written informed consent from all subjects in accordance with the Declaration of Helsinki. The protocols for the use of human blood (permit no. 657-2008) and human wound materials (708-01, 509-01 and 762-2013) were approved by the Ethics Committee at Lund University.

## Data Availability

All data are uploaded to the proteomeXchange repository. The data will be made available upon publication.

## Author contributions

M.vd.P., J.P., S.K. and A.S. conceived and designed the experiments. A.S. and K.S. collected the patient samples. M.vd.P., J.C., J.P. and K.S. prepared the samples and did the *in vitro* analyses. S.K. performed the mass spectrometry measurements. M.vd.P. J.C. and S.K. analysed the results. M.vd.P prepared the figures. M.vd.P and A.S. wrote the paper. All authors discussed the results and commented on the manuscript.

## Acknowledgements

This work was supported by grants from Alfred Österlund Foundation, Edvard Welanders Stiftelse and Finsenstiftelsen (Hudfonden), Lars Hiertas Memorial Foundation, LEO Foundation, O.E. and Edla Johanssons Foundation, the Royal Physiographic Society in Lund, Swedish Research Council (project 2017-02341), the Swedish Government Funds for Clinical Research (ALF) and Åke Wibergs Foundation. The funders had no role in study design, data collection and analysis, decision to publish, or preparation of the manuscript.

## Supplementary Material

**Supplementary Dataset 1** All identified peptides in three plasma samples. List of 770 unique peptide sequences identified by LC-MS/MS

**Supplementary Dataset 2** All identified peptides in five acute wound fluids. List of 7809 unique peptide sequences identified by LC-MS/MS.

**Supplementary Dataset 3**. All identified peptides in 6 dressing extracts. List of 10789 unique peptide sequences identified by LC-MS/MS.

**Supplementary Table 1** Summary of the mass spectrometry results for plasma, acute wound fluids and dressing extracts.

**Supplementary Table 2** Identified peptides and average length from five acute wound fluids.

**Supplementary Table 3** Identified peptides and average length from six dressing extracts.

**Supplementary Figure 1** Venn diagrams comparing the numbers of identified proteins and their peptides of each sample in the low and high inflammation group.

