## supplemental tables and figures for "Method development and characterization of the low molecular weight peptidome of human wound fluids"

Table S1 Summary of mass spectrometry results for the three types of samples

| | Peptides | PSM | Proteins | MeOx | NQ |
|---|---|---|---|---|---|
| **CP** | 770 | 2827 | 65 | 126 (4.5%) | 87 (3.1%) |
| **aWF** | 7809 | 47228 | 373 | 10243 (21.7%) | 3009 (6.4%) |
| **Dressing** | 10789 | 52091 | 418 | 5059 (9.7%) | 1774 (3.4%) |

Total numbers of identified unique peptides, peptide spectrum matches (PSM), and unique proteins found in three citrate plasma samples, five acute wound fluids and six dressing extracts. Furthermore, total numbers and percentages of peptides with post translational modifications (MeOx: methionine oxidation; NQ: deamidation) are indicated.

Table S2 Identified unique peptides, proteins and average peptide length from five acute wound fluids

| | WF1 | WF2 | WF3 | WF4 | WF5 |
|---|---|---|---|---|---|
| **Number of peptides** | 3876 | 4271 | 2932 | 2649 | 3435 |
| **Number of proteins** | 188 | 222 | 150 | 143 | 196 |
| **Average length (Da)** | 1414.70 | 1367.43 | 1463.87 | 1516.11 | 1373.36 |
| **Average number of AA** | 12.69 | 12.28 | 13.26 | 13.68 | 12.28 |

AA: amino acids

Table S3 Identified unique peptides, proteins and average peptide length from six dressing extracts

| | Low inflammation | | | High inflammation | | |
|---|---|---|---|---|---|---|
| | **1** | **2** | **3** | **4** | **5** | **6** |
| **Number of peptides** | 3146 | 4863 | 4277 | 5103 | 1412 | 4929 |
| **Number of proteins** | 179 | 228 | 200 | 170 | 88 | 175 |
| **Average length (Da)** | 1548.67 | 1591.19 | 1546.77 | 1589.84 | 1351.38 | 1668.49 |
| **Average number of AA** | 14.33 | 14.52 | 14.13 | 14.44 | 12.59 | 15.29 |

AA: amino acids

Supplementary figure 1: Venn diagrams comparing the numbers of identified proteins and their peptides of each sample in the low (A) and high (B) inflammation group.

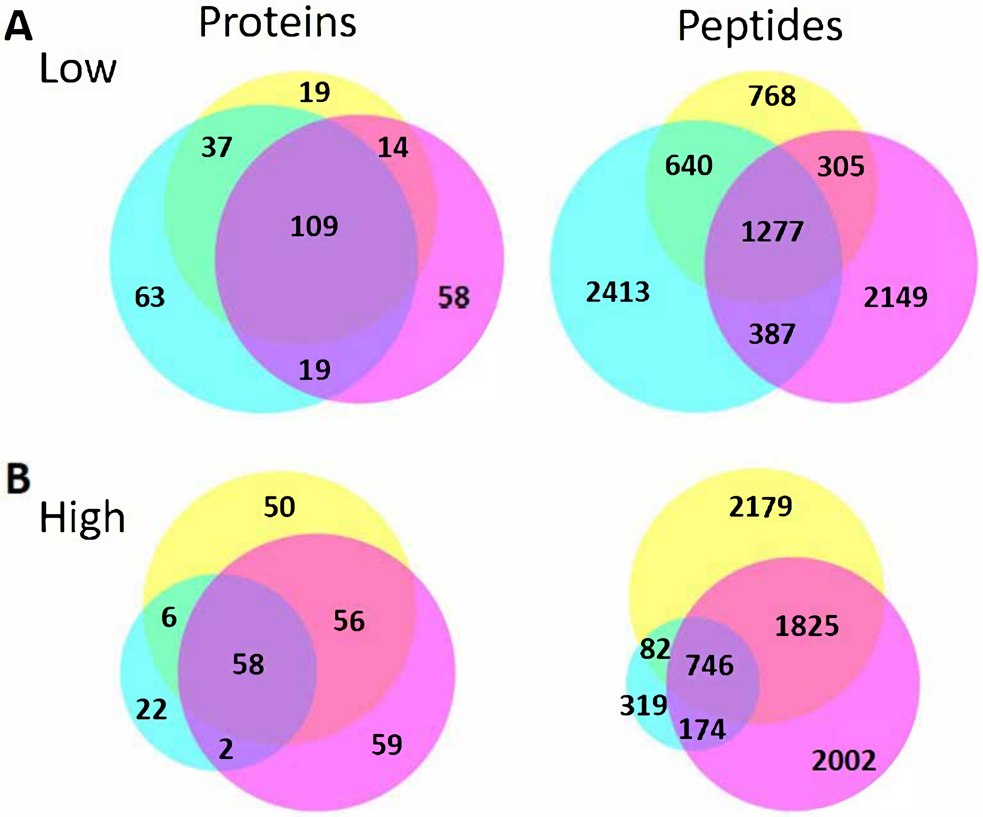
